# Characterization of menopause onset and associated disease risks using large-scale electronic health records

**DOI:** 10.64898/2026.05.08.26352769

**Authors:** Nitya Thakkar, Rajita Patil, Rebecca Levy-Gantt, Yulin Hswen, Monica Agrawal, James Zou, Irene Y. Chen

## Abstract

Menopause affects over one billion women worldwide, yet remains poorly characterized at scale. We apply an ICD-10-based phenotyping algorithm to electronic health records (EHR) from an academic medical center (*n*=33,444 women aged 35–64) and a safety-net hospital system (*n*=7,041), yielding one of the most racially and socioeconomically diverse menopause cohorts in the literature. Structured EHR fields underrepresent symptom burden: only 38.8% of patients had any documented symptom via natural language processing, despite an estimated prevalence of 90%. Adverse pregnancy outcomes were associated with earlier menopause onset after adjustment (*β*=*−*1.21 years, *p*=8.7*×*10*^−^*^45^). Menopausal women showed elevated risk for osteoporosis (hazard ratio of 12.40), rheumatoid arthritis (HR of 2.43), and mental and behavioral disorders (HR 2.38) relative to age-matched men, with divergence at menopause onset. We show that large-scale EHR can characterize menopause at a scale and diversity that prospective enrollment has not achieved.

## 1 Introduction

Menopause is a universal biological transition that affects all women who reach midlife, yet its health consequences remain poorly characterized at the population scale. Defined as the permanent cessation of menstruation after twelve consecutive months [1], menopause, which is accompanied by declining estrogen, coincides with a sharply elevated risk for osteoporosis, cardiovascular disease, and other chronic conditions [2–4]. Understanding when this risk begins, how it varies across populations, and how preventive care should be timed requires large-scale, demographically diverse evidence that existing resources have not provided.

Clinical guidelines governing midlife women’s health rest on evidence from a small number of longitudinal prospective cohorts, including the Study of Women’s Health Across the Nation (SWAN) [4], the Nurses’ Health Studies [5, 6], the Multi-Ethnic Study of Atherosclerosis (MESA) [7], the Women’s Health Initiative [8], and UK Biobank analyses [9–11], which collectively established the menopausal transition as a period of accelerated cardiovascular, metabolic, and skeletal risk (Supplementary Table S1). However, these studies were largely racially homogeneous: the Nurses’ Health Study was 97% White [5], the UK Biobank is over 94% White European and by its own documentation acknowledges limited capacity to study disease risk across ethnic groups [12], and a 2023 re-analysis of SWAN showed that failure to account for weathering [13] caused systematic underenrollment of Black and Hispanic women, masking up to 1.2 years of earlier menopause onset in those groups [14]. Prospective cohort enrollment also tends to recruit healthier and more health-engaged participants, limiting the generalizability of these findings to the broader population of midlife women. Together, this means that large-scale, diverse, electronic helath record (EHR)-based studies of menopausal health outcomes in the United States are essentially absent from the literature, underscoring the need for more scalable, real-world data approaches.

A compounding problem is that menopause is poorly captured even in routine clinical data. Vasomotor symptoms affect an estimated 60–80% of women during the menopausal transition [15], yet in one study, only 23% of women with self-reported vasomotor symptoms had any corresponding documentation in structured EHR fields [16]. Menopause-related International Classification of Diseases, Tenth Revision (ICD-10) codes are applied inconsistently even when symptoms are clinically present [17], in part because the clinical workforce remains ill-equipped to address menopause. A 2022 national survey of 99 Obstetrics and Gynecology residency program directors found that only 31.3% of programs have any dedicated menopause curriculum, and program directors rated the effectiveness of their programs’ menopause training below 4 out of 10 [18]. The result is a systematic measurement gap, where menopause’s clinical burden goes underdocumented even in patients who are actively receiving care.

Beyond the measurement gap, the full breadth of menopause’s health consequences remains undercharacterized. Women in menopause face substantially elevated risk across cardiovascular, skeletal, metabolic, autoimmune, and neurological conditions [2, 19–22], and earlier menopause itself is associated with substantially elevated risks of many chronic diseases and all-cause mortality [7, 9, 11, 22–24]. Capturing the timing and symptomatology of menopause with precision is therefore critical for understanding downstream disease risk. A parallel body of literature has connected adverse pregnancy outcomes (APOs) directly to long-term cardiometabolic risk, with large-scale analyses showing associations between preeclampsia and subsequent cardiovascular disease, preterm birth and all-cause mortality, and gestational diabetes and incident type 2 diabetes persisting decades later [25]. However, the relationship between these earlier reproductive events and menopause timing remains relatively unstudied.

In this work, we address three questions that existing cohorts have been unable to answer at scale (Fig. 1a): What are the clinical symptoms of menopause, and how well do structured medical records capture menopause symptoms? Can obstetric history predict accelerated reproductive aging? Do disease trajectories following menopause onset diverge from those of age-matched men, who do not undergo an analogous hormonal transition? EHR data offer a path to answering these questions, as they capture populations across a full socioeconomic spectrum without active enrollment, offering substantially greater scale and demographic diversity than prospective enrollment. We apply an ICD-10-based phenotyping algorithm to two independent health systems, University of California, San Francisco (UCSF) Health (*n*=33,444) and data collected by the San Francisco Department of Public Health (SFDPH) including safety-net hospitals Zuckerberg San Francisco General and Laguna Honda Hospital (*n*=7,041), yielding one of the most racially and socioeconomically diverse menopause cohorts in the literature (47.9% White at UCSF and 16.9% White at SFDPH), with replication across demographically distinct settings serving as a test of generalizability. In the UCSF cohort, we deploy a two-stage natural language processing (NLP) pipeline using keyword extraction and large language model (LLM)-based context classification to characterize menopause symptom burden across 13 categories from unstructured clinical notes.

**Figure 1:**
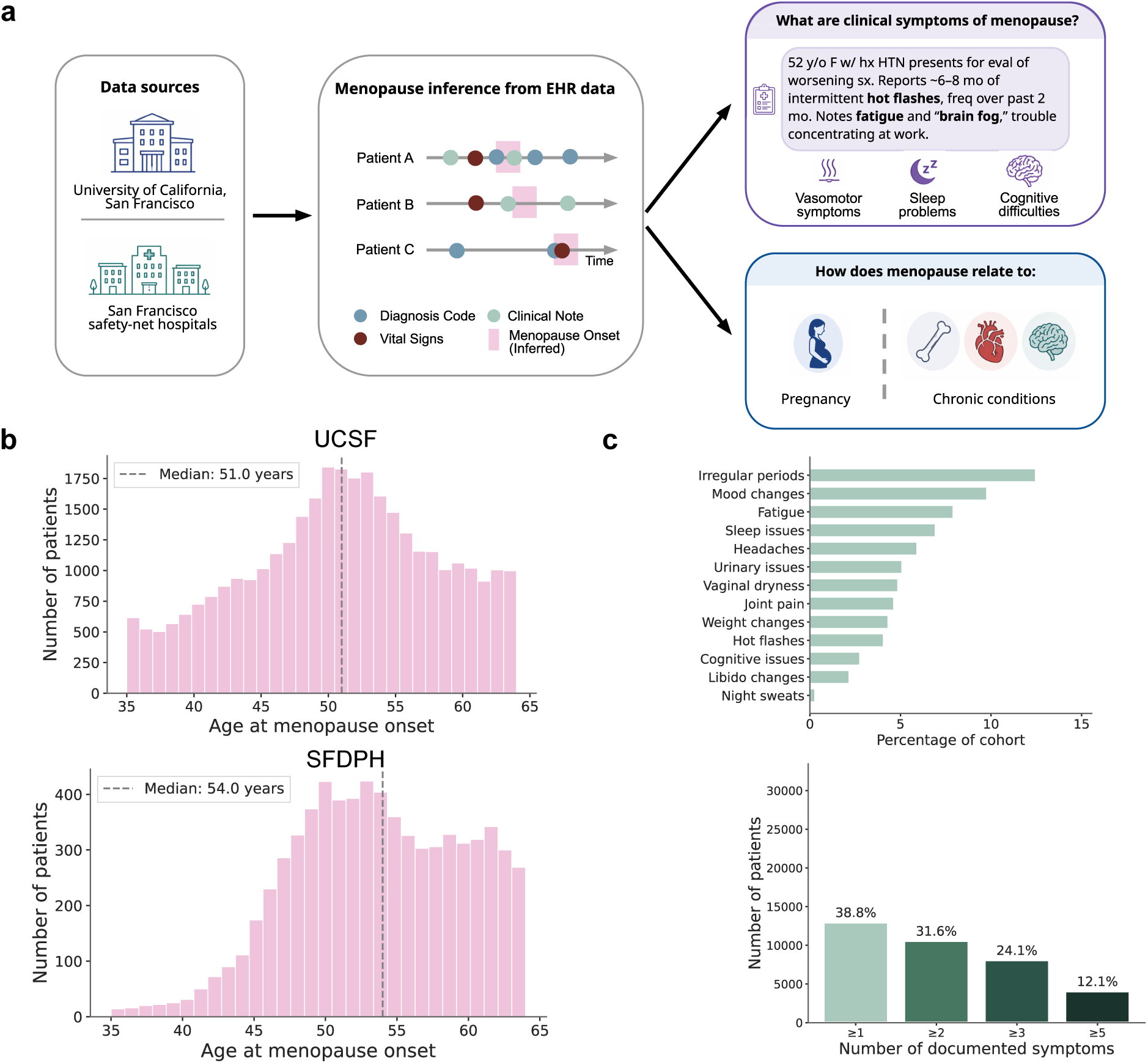
**a.** EHR data from UCSF Health and the SFDPH safety-net system were used to identify menopausal cohorts via ICD-10-based phenotyping, with menopause onset inferred from the earliest qualifying diagnosis code. For patients with clinical notes within ±5 years of menopause onset, we performed NLP-based symptom extraction. We also used extracted menopause data from EHR to understand how menopause onset is related to pregnancy and chronic conditions. **b.** Distribution of age at menopause onset in the UCSF cohort (top; *n*=33,444; median 51 years, IQR 46–56, top) and SFDPH cohort (bottom; *n*=7,041; median 54 years, IQR 49–59, bottom). **c.** Top, prevalence of 13 menopause-related symptom categories extracted from clinical notes via a two-stage NLP pipeline, with irregular periods being the most commonly documented. Bottom, symptom burden distribution across menopausal cohort, with only 38.8% of patients having at least one symptom documented in EHR.

We find that structured EHR fields dramatically underrepresent menopause symptom burden, with only 38.8% of patients having any documented symptoms despite an estimated population prevalence of 90% of women experiencing at least one symptom [26], and that vasomotor symptoms in particular are nearly absent from clinical notes. We show that every APO type examined is associated with earlier menopause onset, with a significant monotonic dose-response relationship, consistent with cumulative obstetric stress accelerating ovarian aging [27, 28]. Finally, we find that women across both cohorts show strikingly elevated risk for osteoporosis (HR 12.40), rheumatoid arthritis (HR 2.43), and mental and behavioral disorders (HR 2.38) relative to age-matched men, with disease trajectories diverging near the time of menopause onset. Together, these findings demonstrate that large-scale EHR analysis combined with NLP can enable more comprehensive and representative characterization of menopause and its associated health outcomes across diverse patient populations.

## 2 Results

### 2.1 Cohort characterization and symptom documentation gap

Menopause onset was inferred from the earliest qualifying ICD-10 code in an eligible encounter (2015-2024), using a clinician-validated code set from prior work [17] capturing symptomatic and asymptomatic menopausal states, premature menopause, and menopausal hormone therapy. Patients were included if assigned female sex at birth and aged 35–64 to capture premature menopause while avoiding Medicare-related utilization artifacts.

Applying this inclusion criterion to the UCSF EHR yielded a menopausal cohort of *n*=33,444 women with a median age at onset of 51 years (Fig. 1b top). The established US mean age at natural menopause is approximately 50 years [1, 29], consistent with our median of 51 years (mean: 50.9 years). Applying the same inclusion criterion to an independent cohort of patients from the SFDPH EHR, a safety-net population that is demographically and socioeconomically distinct from the primary UCSF cohort (see Supplementary Table S2), yielded a cohort of *n*=7,041. In this cohort, the median menopause age was 54 years (mean: 53.7 years), approximately three years higher than in the UCSF cohort (Fig. 1b bottom); this higher median age likely reflects patterns of delayed engagement with specialty care in this population.

Menopause timing differed significantly by race and ethnicity after adjustment for confounding variables, although the patterns were site-specific. In the UCSF data (Supplementary Fig. S1a), we observed that Asian women reached menopause a mean of 2.6 years earlier than White women (White mean: 51.7 years) and Latinx women a mean of 2.0 years earlier than White women (both *p <* 0.05; FDR-adjusted). While Black women reached menopause a mean of 0.8 years earlier than White women, this was not statistically significant after adjusting for confounders. However, in the SFDPH data (Supplementary Fig. S1b), we observed that Asian women (Asian mean: 56.0 years) reached menopause significantly later than White patients, by a mean of 2.8 years, and Black patients, by a mean of 3.3 years (both *p <* 0.05; FDR-adjusted). In both the UCSF (Supplementary Fig. S1c) and SFDPH cohorts (Supplementary Fig. S1d), patients of Hispanic/Latino ethnicity experienced menopause onset significantly earlier than patients not of Hispanic/Latino ethnicity (UCSF: mean 1.3 years earlier, SFDPH: mean 4.1 years earlier).

Furthermore, we found that structured EHR fields captured only a fraction of menopause symptom burden in this cohort. A recent study of 1,510 women by the American Association of Retired Persons found that 90% of women experienced at least one symptom, with an average of 5 symptoms per woman [26]. However, LLM-based extraction from unstructured clinical notes identified at least one documented symptom in only 38.8% of patients (*n*=12,969), with irregular periods (24.9%), mood changes (19.5%), fatigue (15.8%), sleep issues (13.8%), and headaches (11.8%) being the five most prevalent (Fig. 1c top). Notably, hot flashes, widely considered the most common menopause symptom, appeared in just 8.1% of records, and night sweats in 0.5%, despite their well-documented prevalence in other survey studies [15, 26, 30]. We hypothesize these numbers reflect menopause symptoms that are experienced but not elicited, as clinicians (many of whom are likely underprepared to manage menopause [18]) may not be asking patients about them consistently. Among symptomatic patients, reporting of multiple symptoms was common, with 31.6% of the full cohort having at least 2 symptoms, 24.1% having at least 3, and 12.1% having at least 5 (Fig. 1c bottom).

We provide a full breakdown of the LLM categorization statistics in Supplementary Table S3, and symptom co-occurrence patterns are provided in Supplementary Fig. S2. Notably, the five strongest pairwise associations were concentrated among neuropsychological and somatic symptoms (mood changes, fatigue, sleep issues, headaches), while vasomotor symptoms (hot flashes, night sweats) and genitourinary symptoms (vaginal dryness, urinary issues) showed little co-occurrence with other symptom categories.

### 2.2 Adverse pregnancy outcomes predict earlier menopause onset

Adverse pregnancy outcomes (APOs), defined here as gestational diabetes, preterm birth, pre-eclampsia, and gestational hypertension, determined via ICD-10 codes, are well-established markers of long-term cardiometabolic risk, yet whether they accelerate ovarian aging and link obstetric history to earlier menopause onset has received minimal study. A 2023 study found that certain APOs, such as preterm birth with spontaneous labor, were associated with younger age at natural menopause, and that other APOs, such as gestational diabetes and hypertensive disorders of pregnancy, were associated with older age at natural menopause [6]. In our data, all four APO types were associated with younger menopause age, including gestational diabetes.

We analyzed all women in the UCSF menopausal cohort with documented pregnancy history (*n*=9,717). This subgroup had a median menopause onset age of 51.2 (IQR 46.0–56.1) years, a mean age at delivery (across all recorded births) of 34.14 years, and a similar racial demographic breakdown to the full cohort. For these patients, each APO type examined was significantly associated with earlier menopause onset in unadjusted comparisons (Mann-Whitney *p <* 0.0001; Fig. 2a). The largest unadjusted gap was observed for gestational diabetes (median onset 43.8 years, IQR 40.2–48.2) relative to women with no documented complications (median 53.2 years, IQR 49.1–57.6). After full confounder adjustment, each APO type remained significantly associated with earlier menopause onset (pre-eclampsia: *p <* 0.0001; gestational diabetes: *p <* 0.0001; gestational hypertension: *p <* 0.0001; preterm birth: *p <* 0.0001; all FDR-adjusted), with any APO versus no complications associated with 1.2 years earlier onset (*β* = −1.21 years, 95% confidence interval (CI) −1.37 to −1.04, *p* = 8.7 × 10*^−^*^45^). To address the concern that women with APO histories engage more frequently with the healthcare system and therefore receive earlier menopause codes, all models were adjusted for healthcare utilization as a covariate.

**Figure 2:**
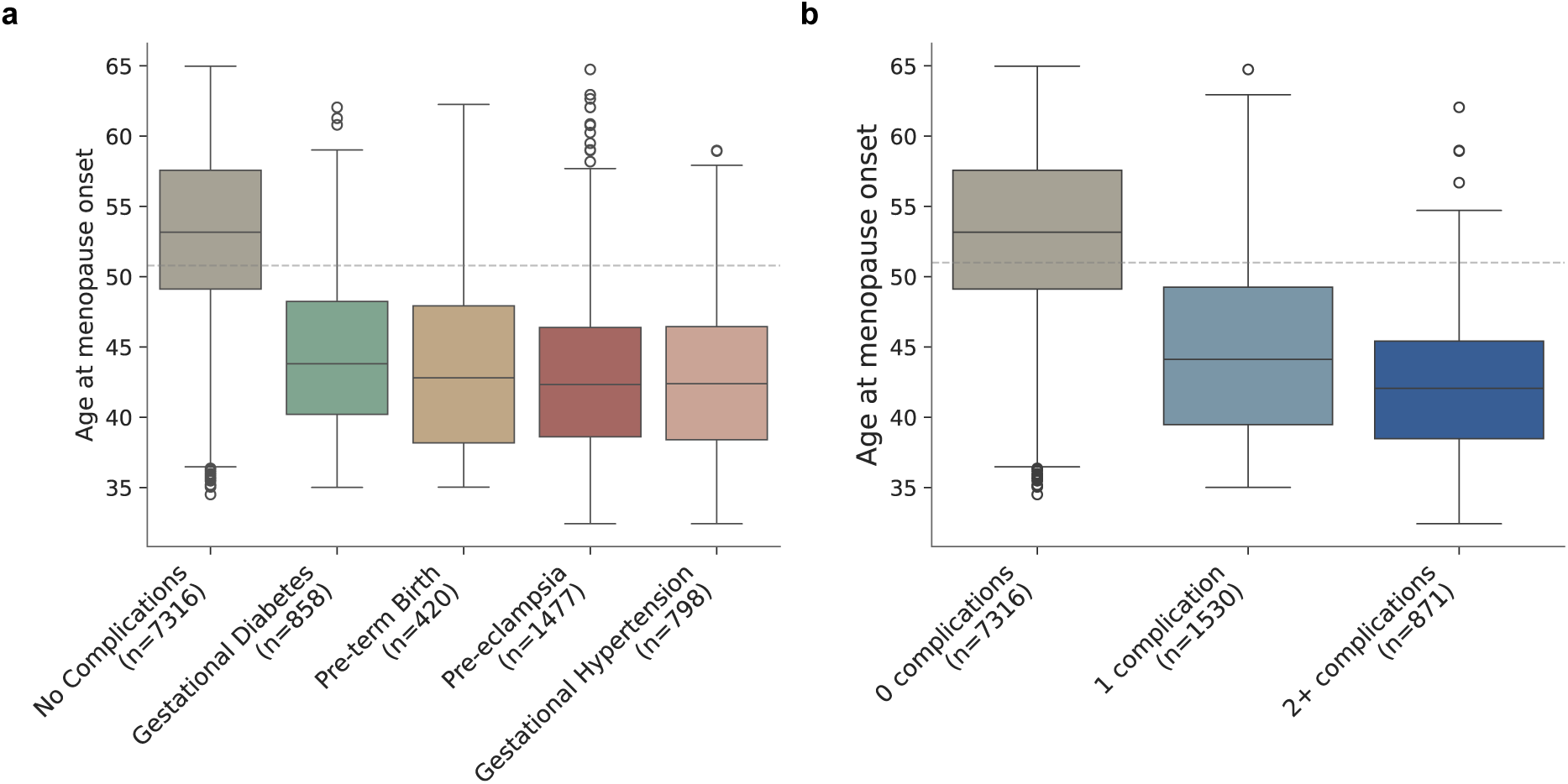
**a.** Women with gestational diabetes (median: 43.8 years), preterm birth (median: 42.8 years), preeclampsia (median: 42.4 years), and gestational hypertension (median: 42.3 years) reached menopause significantly earlier than women with no documented complications (median: 53.2 years). Unadjusted median differences are shown in the figure, and unadjusted p-values are computed with the two-sided Mann-Whitney U test and reported in the main text. Data are presented as median with interquartile range. The gray dashed line represents the median age of menopause onset in the UCSF cohort (51 years). **b.** Women with 1 (median: 44.1 years) and 2+ (median: 42.1 years) APOs reached menopause progressively earlier, consistent with a dose-response relationship between cumulative obstetric stress and accelerated ovarian aging. The monotonic decreasing trend across ordered burden groups was significant (Jonckheere-Terpstra: *J* = −0.43, *p <* 0.0001).

The relationship between cumulative APO burden and menopause timing showed a significant monotonic dose-response (Fig. 2b). Women with 1 and 2+ APOs reached menopause progressively earlier in adjusted analyses (1 vs. 0 APOs: *β* = −1.14 years, 95% CI −1.32 to −0.96, *p* = 1.6 × 10*^−^*^33^; 2+ vs. 0 APOs: *β* = −1.32 years, 95% CI −1.60 to −1.05, *p* = 7.4 × 10*^−^*^21^). A Jonckheere-Terpstra test confirmed a significant monotonic decreasing trend across ordered burden groups (*J* = −0.43, *p <* 0.0001), consistent with a dose-response relationship between cumulative APO burden and accelerated reproductive aging. While the adjusted effect sizes are modest in absolute terms, they are consistent with the 1–3 year effects reported in prior prospective cohorts [6]. Furthermore, these findings are clinically meaningful at the population scale, as a one-year advancement in menopause onset can correspond to earlier onset of accelerated disease risk for thousands of women annually.

### 2.3 Disease trajectories diverge from age-matched men after menopause onset

We next characterized how the menopausal transition marks the onset of diverging disease trajectories. Prior studies have largely treated menopause as a binary cross-sectional exposure, and while prospective work has linked reproductive period length to dementia risk [22] and premature menopause to a 55% higher cardiovascular disease risk [23], no study has characterized simultaneous trajectory divergence across multiple organ systems relative to menopause onset in a diverse real-world population.

We fit risk-adjusted Cox proportional hazards models to our menopausal cohort, excluding those with prior disease history (Fig. 3a). Women were matched to men on birth year, with calendar year, healthcare utilization, demographics, and socioeconomic confounders adjusted as covariates in the Cox models. Osteoporosis showed by far the largest effect, with women more than twelve times as likely to receive an incident diagnosis as age-matched men (hazard ratio (HR) 12.40, 95% CI 10.55 to 14.56, *p* = 1.3×10*^−^*^206^). Atherosclerosis (HR 3.96, 95% CI 3.18 to 4.93, *p* = 1.9×10*^−^*^34^), rheumatoid arthritis (HR 2.43, 95% CI 1.70 to 3.50, *p* = 1.4×10*^−^*^6^), mental/behavioral disorders (HR 2.38, 95% CI 2.22 to 2.56, *p* = 4.7×10*^−^*^126^), and lupus (HR 2.61, 95% CI 1.46 to 4.69, *p* = 0.0013) were all significantly elevated in women. Alzheimer’s disease showed an elevated hazard (HR 1.80, 95% CI 1.04 to 3.09, *p* = 0.035), but should be interpreted cautiously given the small number of cases and marginal p-value; this finding did not replicate in the SFDPH cohort (HR 1.15, *p* = 0.80). Women had significantly lower hazard than men for several cardiometabolic conditions: heart failure (HR 0.33, 95% CI 0.28 to 0.39, *p* = 9.6×10*^−^*^43^), stroke (HR 0.79, 95% CI 0.69 to 0.91, *p* = 0.0012), hypertension (HR 0.69, 95% CI 0.65 to 0.74, *p* = 1.4×10*^−^*^32^), ischemic heart disease (HR 0.87, 95% CI 0.80 to 0.93, *p* = 1.5×10*^−^*^4^), and type 2 diabetes (HR 0.45, 95% CI 0.41 to 0.50, *p* = 3.3×10*^−^*^48^). Women also showed significantly lower hazard for Parkinson’s disease (HR 0.30, 95% CI 0.20 to 0.45, *p* = 4.8×10*^−^*^9^). All p-values are false discovery rate (FDR)-adjusted, and hazard ratios are included in Supplementary Table S4. To illustrate some of these trends, we visualized cumulative prevalence curves across the ±10 years surrounding menopause onset for osteoporosis, rheumatoid arthritis, and mental/behavioral disorders. Cumulative prevalence curves showed upward trajectories for all three conditions, with divergence beginning near the time of menopause onset (Fig. 3b). The cumulative prevalence curves are intended to illustrate trajectory divergence relative to menopause onset, not to compare absolute disease burden between women and men, whose pre-menopause baseline rates differ.

**Figure 3:**
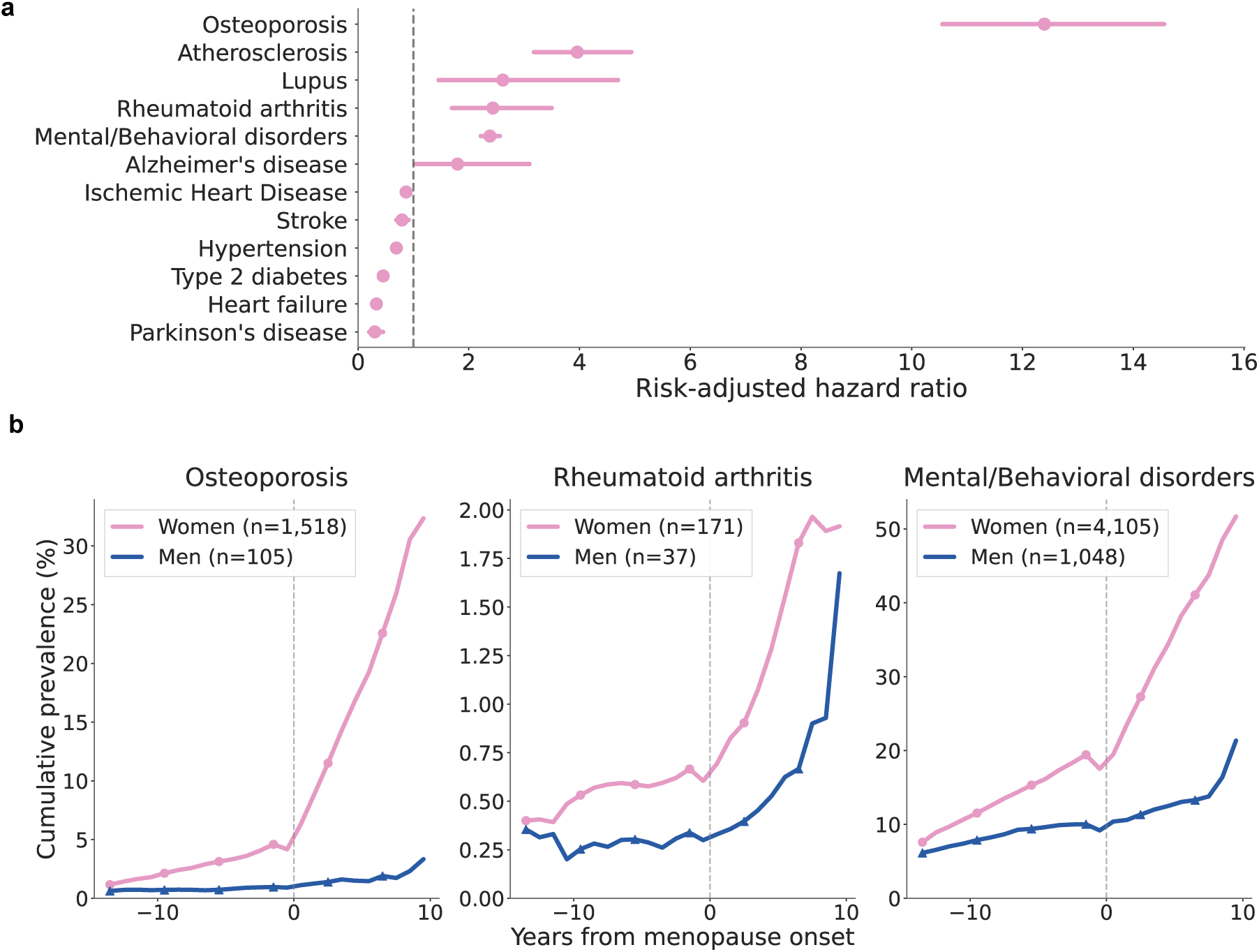
**a.** Risk-adjusted hazard ratios for 12 incident conditions in women after menopause onset, compared to age-matched men, adjusted for birth year, healthcare utilization, demographic, and socioeconomic confounders. Points to the right of the dashed line (HR=1) indicate elevated risk in women after menopause onset compared to men. Osteoporosis showed the largest sex-specific elevation (HR 12.40). **b.** Cumulative prevalence curves across the ±10 years surrounding menopause onset for osteoporosis, rheumatoid arthritis, and mental/behavioral disorders, comparing women (pink) to age-matched men (blue). Prevalence visually accelerates in women following menopause onset for all three conditions.

### 2.4 Disease trajectory patterns replicate in an independent safety-net population

Risk-adjusted Cox proportional hazards models, fit to our SFDPH menopausal cohort (excluding those with prior disease history), matched to men on birth year and adjusted for calendar year, healthcare utilization, demographics and socioeconomic confounders, revealed broadly consistent trends (Fig. 4a). Osteoporosis again showed by far the largest signal, with women more than ten times as likely to receive an incident diagnosis as age-matched men (HR 10.83, 95% CI 8.59 to 13.66, *p* = 3.2×10*^−^*^90^). Hypertension (HR 1.14, 95% CI 1.03 to 1.25, *p* = 0.0091), rheumatoid arthritis (HR 4.09, 95% CI 2.33 to 7.17, *p* = 9.0×10*^−^*^7^), mental/behavioral disorders (HR 2.56, 95% CI 2.32 to 2.84, *p* = 4.4×10*^−^*^75^), and lupus (HR 4.37, 95% CI 1.40 to 13.61, *p* = 0.011) were all significantly elevated in women. Alzheimer’s disease showed a non-significant result (HR 1.15, 95% CI 0.39 to 3.41, *p* = 0.80). Women had significantly lower hazard than men for several cardiometabolic conditions: heart failure (HR 0.65, 95% CI 0.53 to 0.80, *p* = 5.9×10*^−^*^5^), atherosclerosis (HR 0.65, 95% CI 0.49 to 0.85, *p* = 0.0018), ischemic heart disease (HR 0.74, 95% CI 0.62 to 0.88, *p* = 5.0×10*^−^*^4^), stroke (HR 0.78, 95% CI 0.64 to 0.95, *p* = 0.013), and type 2 diabetes (HR 0.73, 95% CI 0.63 to 0.84, *p* = 9.0×10*^−^*^6^). Women also showed significantly lower hazard for Parkinson’s disease (HR 0.38, 95% CI 0.16 to 0.90, *p* = 0.028).

**Figure 4:**
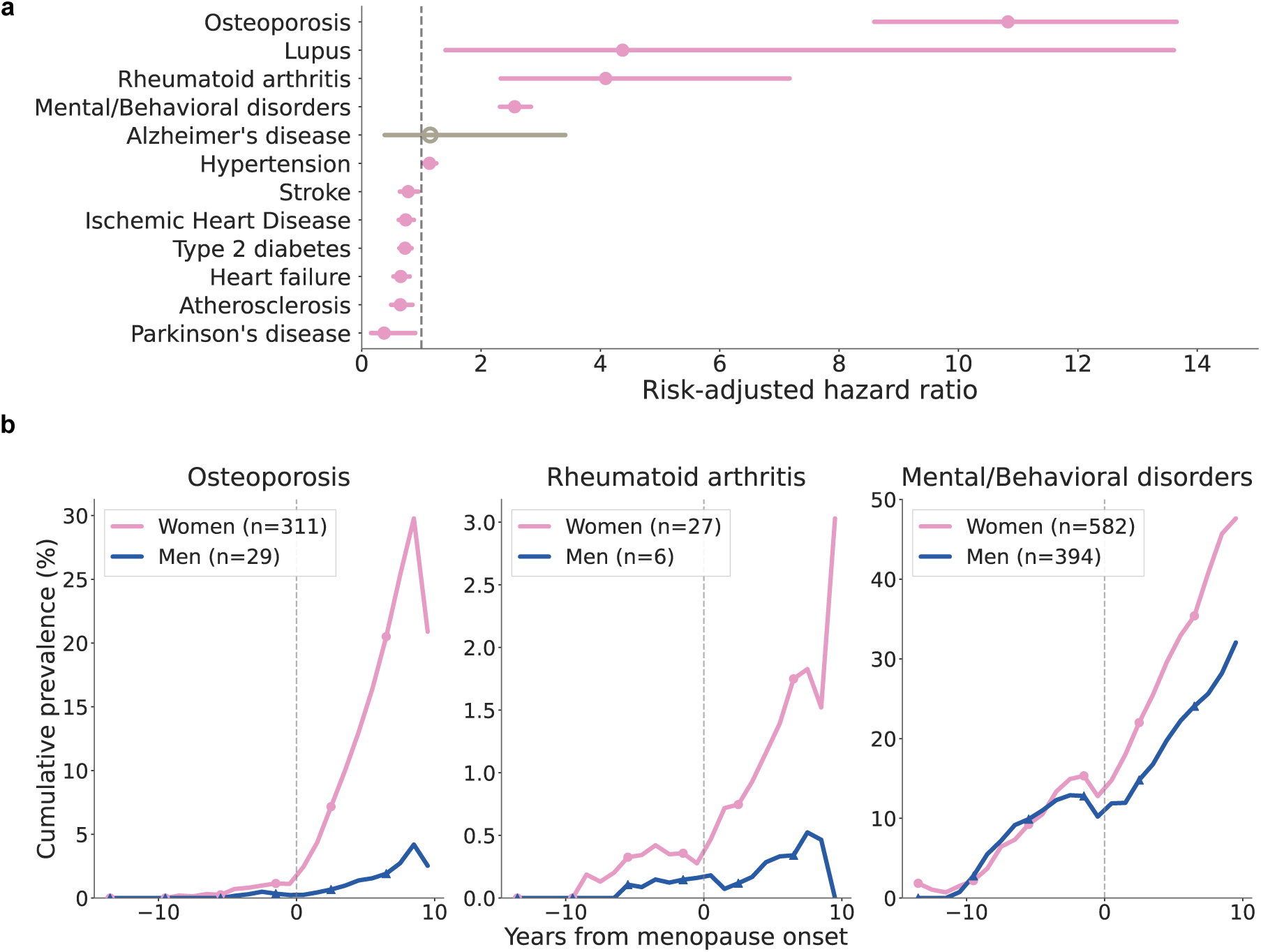
**a.** Risk-adjusted hazard ratios for 12 incident conditions in the SFDPH cohort, comparing women after menopause onset to age-matched men, adjusted for birth year, healthcare utilization, demographic, and socioeconomic confounders. Points to the right of the dashed line (HR=1) indicate elevated risk in women after menopause onset compared to men. Hazard ratio estimates between SFDPH and UCSF cohorts were highly correlated (Pearson *r*=0.80, *p*=1.8×10*^−^*^3^). **b.** Cumulative prevalence curves for the SFDPH cohort across the ±10 years surrounding menopause onset for osteoporosis, rheumatoid arthritis, and mental/behavioral disorders, comparing women (pink) to age-matched men (blue). Divergence after menopause onset replicated across all conditions.

We found that risk-adjusted hazard ratio estimates across 12 conditions were highly correlated with the primary UCSF cohort (Pearson *r*=0.80, *p*=1.8×10*^−^*^3^). The two notable discrepancies between cohorts were hypertension and atherosclerosis. In the UCSF cohort, hypertension showed a significantly lower hazard in women (HR 0.69), while in SFDPH, women showed a higher risk (HR 1.14). This reversal is consistent with the elevated baseline hypertension prevalence in the safety-net population, where the male comparison cohort has a higher pre-existing burden. Atherosclerosis showed elevated risk in UCSF women (HR 3.96) but a protective effect in SFDPH women (HR 0.65), potentially reflecting ICD-10 coding practices: UCSF’s academic cardiology infrastructure may document atherosclerosis more consistently than a safety-net setting, inflating apparent incidence in women relative to men. These discrepancies highlight the sensitivity of ICD-10-based incidence estimates and underscore the importance of interpreting hazard ratios in the context of site-specific documentation practices and patient population characteristics. All SFDPH hazard ratios, alongside UCSF hazard ratios, are in Supplementary Table S4. Finally, we observed that disease divergence patterns, visualized as cumulative prevalence curves, replicated across osteoporosis, rheumatoid arthritis, and mental/behavioral disorders (Fig. 4b).

## 3 Discussion

We show that large-scale EHR analysis, combined with NLP techniques, can characterize menopause timing, symptom burden, and downstream disease risk at a scale and diversity that prospective cohort enrollment has not achieved. Adverse pregnancy outcomes were, after adjustment, associated with 1.2 years earlier menopause onset with a significant monotonic dose-response. We further find that menopausal women across the socioeconomically and demographically diverse cohorts face substantially elevated risk for osteoporosis, rheumatoid arthritis, and mental and behavioral disorders relative to age-matched men, with disease trajectories diverging near the time of menopause onset. Many of these trends are consistent with prior work on atherosclerosis [31], rheumatoid arthritis [21], mental health [32], lupus [33], and Alzheimer’s disease [11, 22]. While women’s relative hazard for cardiovascular conditions is lower than men’s following menopause onset, we observed that the absolute risk of these conditions nonetheless increases after the menopausal transition. Gestational diabetes was associated with earlier rather than later menopause in our cohort, which diverges from prior prospective work [6] and likely reflects differences in ascertainment methodology and cohort composition. Prior prospective studies were predominantly White, whereas our cohort is substantially more racially and socioeconomically diverse, and ovarian aging trajectories are known to vary across racial groups [34]. Our ICD-10-based design also likely captures women with more severe presentations of gestational diabetes who subsequently engaged with menopause specialty care. The dose-response pattern we observe is biologically consistent with cumulative oxidative stress on ovarian reserve, as each APO type is independently associated with elevated systemic oxidative stress during pregnancy [27], and oxidative stress is a well-established driver of accelerated follicular depletion [28], such that each additional adverse pregnancy outcome likely adds to the ovarian burden and advances menopause timing.

Overall, our findings suggest that menopause represents a critical inflection point in long-term health trajectories, where biological, reproductive, and systemic risk factors converge. Improved identification of menopause timing, therefore, has the potential to enable earlier and more targeted preventive care, particularly when integrated with longitudinal reproductive history. Obstetric history, which is already routinely documented in the EHR, should be incorporated into midlife women’s health risk stratification. Women with APOs may warrant earlier initiation of osteoporosis screening, cardiovascular risk assessment, and mental health monitoring.

The striking underrepresentation of menopause symptoms in EHR notes (38.8% documentation despite 90% population prevalence) similarly points to a concrete clinical opportunity. Structured symptom checklists at relevant encounter types, such as gynecology visits and primary care preventive visits, would substantially improve symptom documentation without requiring new infrastructure. This underrepresentation reflects not just a documentation practice problem but a code granularity problem. For example, N95.1 (Menopausal and Female Climacteric States) conflates symptomatic and asymptomatic menopause, and a clinician may apply it both to a patient with severe hot flashes and to one with no symptoms, producing ambiguous structured data. The ICD-10 system, as currently structured, lacks the specificity needed to distinguish symptom burden from menopausal status, limiting the utility of structured EHR fields for menopause research regardless of clinician behavior. Our findings also suggest that menopause symptom burden is not only underdocumented but inherently multidimensional. Among symptomatic patients, co-occurring symptoms were common, with nearly a quarter of the cohort experiencing three or more documented symptoms. Co-occurrence was strongest among neuropsychological and somatic symptoms (mood changes, fatigue, sleep disturbance, and headache), while vasomotor symptoms were less prominently represented in clinical documentation despite their high population prevalence. Menopause is not defined by a single dominant symptom but by an interconnected symptom network that current documentation practices fail to capture, and failure to represent this complexity in the EHR may contribute to under-recognition of the full clinical burden experienced by patients.

Menopausal hormone therapy (MHT), formerly referred to as hormone replacement therapy, represents another area where our findings reveal a gap between documented need and delivered care. MHT has received renewed attention following the Women’s Health Initiative’s long-term follow-up, which found no significant increase in all-cause or cause-specific mortality with MHT use [35], reducing the concerns that sharply reduced its use in the early 2000s. Yet in our cohorts, MHT use was strikingly low: 29.4% of UCSF patients and just 3.0% of SFDPH patients at any point. The more than tenfold gap between sites reflects a structural equity problem in access to hormonal intervention, not a difference in disease burden. The observational design of this study does not allow for causal inference about the effects of MHT on disease outcomes, and residual confounding from unmeasured variables remains possible, but the disparity between these statistics and the disease burden we document suggests a large gap between the population that might benefit from preventive hormonal intervention and the population currently receiving it.

Several important limitations warrant consideration. Selection bias is a fundamental constraint of ICD-10-based cohort construction, as patients with documented menopause are, by definition, patients who engaged with the healthcare system and received a diagnosis code, meaning they likely experience more health problems and interact more frequently with clinical care than the average menopausal woman. This bias affects both whom we study and what we measure, and may inflate both symptom and disease prevalence estimates relative to the general population [36]. We also assume that ICD-10 codes reliably distinguish natural from surgical or medically induced menopause, and possible misclassification of menopause type could confound the timing and disease risk analyses. The reversal of the Asian-White menopause timing difference across the two cohorts is most plausibly a measurement artifact of this same ascertainment process, as ICD-10-based menopause coding requires a clinical encounter in which menopause is documented, introducing selection bias from differential healthcare access [16, 17], and the SFDPH safety-net population likely has delayed care-seeking that inflates apparent menopause age. The residual racial differences within each site, after adjustment for socioeconomic confounders, may additionally reflect genuine biological effects of socioeconomic factors on ovarian aging [34]. Pregnancy history and APO data were unavailable in the SFDPH dataset, limiting the APO analysis to the UCSF cohort and preventing replication of the dose-response finding in the independent safety-net population. The SFDPH cohort also did not include clinical notes, so symptom-level findings could not be replicated there, and the full extent of documentation failure across safety-net settings remains unknown.

Finally, both health systems studied are in the San Francisco Bay Area, and while the SFDPH safety-net population considerably extends demographic diversity, generalization to other geographic and socioeconomic contexts remains to be established. Beyond the two cohorts studied here, the National Institutes of Health’s All of Us Research Program [37] offers a promising extension, as its enrollment was designed to prioritize populations historically underrepresented in biomedical research and includes a higher proportion of women than most existing biobanks [38]. Furthermore, social media and online patient communities have emerged as a complementary data source where women discuss menopause precisely because clinical settings have not provided adequate space for it [39, 40], though these carry their own limitations, including non-representative sampling and susceptibility to misinformation [41].

The documentation failures in our cohort are consistent with the well-documented gap in clinical preparedness for menopause care [18]. Retraining the clinical workforce is important but costly and slow. Computational tools embedded in clinical workflows, such as those that prompt symptom assessment and flag high-risk patients based on obstetric history, represent a more immediately scalable path to closing this gap. The divide between what these data reveal and what happens in the average clinical encounter is not a knowledge problem but a structural one, and it is a problem that computational tools, better training, and more equitable access to intervention can begin to close.

## 4 Methods

### 4.1 Study populations

The UCSF de-identified Clinical Data Warehouse (DeID CDW) integrates structured EHR data and deidentified clinical notes from UCSF Health facilities, including UCSF Medical Center, UCSF Benioff Children’s Hospitals, UCSF Dental Center, and Langley Porter Psychiatric Hospital and Clinics, with data through July 2025 [42]. The San Francisco Department of Public Health (SFDPH) DeID CDW integrates structured EHR data through December 2024, without clinical notes, from safety-net systems including: Zuckerberg San Francisco General Hospital and Trauma Center, Laguna Honda Hospital, Population Health Division, Behavioral Health Services, and ambulatory care areas [43]. The SFDPH dataset excludes patients who received Maternal, Child, and Adolescent Health (MCAH) services (*n*=14,043). Both datasets are de-identified and do not require review by an Institutional Review Board (IRB).

### 4.2 Cohort construction

We identified women with at least one menopause-related ICD-10 code documented during an eligible encounter from January 1, 2015 to December 31, 2024: Z78.0 (asymptomatic menopausal state), N95.1 (menopausal and female climacteric states), N95.8 (other specified menopausal and perimenopausal disorders), N95.9 (unspecified menopausal and perimenopausal disorder), E28.310 (symptomatic premature menopause), E28.319 (asymptomatic premature menopause), E28.39 (other primary ovarian failure), and Z79.890 (hormone replacement therapy). This code set was drawn from prior literature [17] and validated by a board-certified Obstetrician and Gynecologist (OB/GYN). We added Z79.890 (the ICD-10 code designated ‘hormone replacement therapy’) based on clinical consultation with an OB/GYN. Patients were required to be assigned female sex at birth and aged 35–64 years at the qualifying encounter. The lower bound of 35 years was chosen to exclude the age range most commonly associated with premature ovarian insufficiency, while still capturing premature and early-onset menopause. The upper bound of 64 avoids Medicare-related changes in healthcare utilization.

For each patient, we extracted demographics such as race (categorized as White, Asian, Black, Latinx, or Other) and ethnicity (Hispanic/Latino or not Hispanic/Latino). We also extracted all proxies for socioeconomic status available in the data, including preferred language, marital status, postal code, smoking status, body mass index (BMI; last recorded value on or before menopause onset date, imputed using median for those missing), menopausal hormone therapy use (defined by ICD-10 Z79.890 presence), and healthcare utilization (measured as unique encounter-days per year). For all analyses, we adjust for these confounding variables. We provide a full overview of covariates collected in Supplementary Table S2. We excluded lab measurements, specifically follicle-stimulating hormone, estradiol, luteinizing hormone, and anti-Müllerian hormone, due to high missingness in our cohort.

Differences in age at menopause onset by race and ethnicity were estimated using confounder-adjusted linear regression with age at first qualifying ICD-10 code as the outcome. Models were fit separately for each racial and ethnic group relative to White women as the reference.

### 4.3 Symptom extraction from clinical notes

NLP-based approaches to menopause in clinical settings remain rare. The most directly relevant prior work applied a rule-based NLP system to Veterans Affairs (VA) clinical notes to classify menopausal status, distinguishing pre-from post-menopausal patients, and found that NLP increased the proportion of classifiable patients from 27% to 55% [44]. A separate study applied NLP to online patient community posts to characterize vasomotor symptom burden and management, but relied on social media rather than clinical notes and did not attempt population-level phenotyping [39]. While neither study characterized symptom burden from routine clinical notes at the population scale, recent work has shown that LLMs are even more powerful and accurate EHR symptom extractors than ICD-10 codes in other clinical domains [45].

Symptom burden was characterized from unstructured clinical notes using a two-stage pipeline. In the first stage, all notes for our cohort ±5 years from their menopause onset date were selected. We used regular expression (regex) matching to search for a predefined set of symptoms drawn from the Menopause Rating Scale [46], the Kupperman Menopausal Index [47], and symptom domains documented in SWAN [4]. The 13 categories were: hot flashes, night sweats, irregular periods, vaginal dryness, mood changes, sleep issues, cognitive issues, joint pain, weight changes, fatigue, headaches, urinary issues, and libido changes. An OB/GYN reviewed this symptom list. We provide a complete mapping of keywords used in Supplementary Table S5. In this step, we extracted 88,132 notes belonging to 13,985 menopause patients in our cohort.

In the second stage, symptom-level context classification was performed on the notes flagged as containing at least one of the 13 target symptoms using the GPT-OSS-20B LLM [48]. For each symptom flagged in a note, the model assigned one of seven context labels: current, not mentioned, negated, past, concern, family history, or uncertain. Only mentions classified as current were counted toward prevalence estimates. Symptom prevalence estimates are reported as the proportion of the full UCSF menopausal cohort (*n*=33,444) with at least one current mention of each symptom category; patients lacking any clinical notes in the ±5-year window were counted as having no documented symptoms. The high rate of ‘Not Mentioned’ classifications (Supplementary Table S3) confirms the LLM’s ability to reject regex false positives from the keyword extraction step.

The LLM symptom-level context classification was validated against expert human annotation (blinded to the LLM outputs) of 130 stratified examples across 13 symptom categories (*n*=10 per category). The LLM achieved an overall accuracy of 0.80 and F1 of 0.89 for the current label (precision = 0.91, recall = 0.88), which drove all prevalence estimates reported in this study (see Supplementary Fig. S3). Per-symptom F1 for ‘current’ ranged from 0.67 to 1.00 across symptom categories, with mood changes showing the lowest agreement (F1 = 0.67), likely reflecting clinical ambiguity in mood-related documentation. The ’not mentioned’ label, which captures regex false positives, achieved F1 = 0.80, validating the false positive rejection mechanism of this two-stage pipeline.

### 4.4 Extracting adverse pregnancy outcomes

Adverse pregnancy outcomes (APOs) were identified by ICD-10 codes from women with documented pregnancy history (*n*=9,717): pre-eclampsia (Z87.59, O14, O15), gestational diabetes (O24), gestational hypertension (O13), and preterm birth (O60), following prior work [25]. Women with a history of spontaneous abortion (O03, n=47) or small for gestational age (P05.10, n=9) were not included as APO types due to small sample size; we excluded them from the no-complication reference group. Differences in age at menopause onset across APO groups were assessed using the Kruskal-Wallis test. A Jonckheere-Terpstra test was used to formally test the monotonic trend in menopause age across ordered APO burden groups. For the burden analysis, the APO count was grouped as 0, 1, or 2+ complications. Adjusted effect estimates were obtained from ordinary least squares linear regression with menopause age as the outcome and APO status as the primary predictor. Multiple testing across individual APO types was corrected using Benjamini-Hochberg FDR.

To assess whether the APO-menopause timing association was driven by ascertainment bias, whereby women with APO histories engage more frequently with the healthcare system and therefore receive earlier menopause codes, we adjusted all models for healthcare utilization (unique encounters per year). Additionally, models were adjusted for all the previously described confounding variables, along with parity (number of births) and age at first pregnancy.

### 4.5 Analyzing disease burden

To separate the sex-specific disease burden of the period following menopause onset from the effects of aging, we constructed a male comparison cohort matched to women on birth year (±2 years), with calendar year and healthcare utilization adjusted as covariates in the Cox models. Up to four controls were matched per woman, following prior work [49], with men appearing in multiple women’s eligible pools retained once with the earliest assigned reference date.

Twelve incident outcome conditions were defined by ICD-10 code prefix: atherosclerosis (I70), Alzheimer’s disease (G30), Parkinson’s disease (G20), rheumatoid arthritis (M05, M06), heart failure (I50), hypertension (I10–I13, I15, I16), ischemic heart disease (I20–I25), stroke (I60–I69), type 2 diabetes (E11), osteoporosis (M80, M81), mental and behavioral disorders (F0–F9), and lupus (L93, M32). Patients with a qualifying diagnosis before the reference date were excluded from each respective outcome analysis.

Cox proportional hazards models were fit using the Python lifelines package [50], with time to first incident diagnosis as the outcome and female sex as the main risk factor. All models were adjusted for the previously described confounding variables along with age at reference date and calendar year. Cumulative prevalence curves were computed in annual time bins anchored to the reference date across ±10 years. Multiple testing correction was applied using the Benjamini-Hochberg false discovery rate procedure across the 12 outcome conditions, with a significance threshold of FDR-adjusted *p* ≤ 0.05.

## Data availability

The UCSF DeID CDW [42] and SFDPH DeID CDW [43] datasets are de-identified clinical relational data repositories available to UCSF researchers through the UCSF Information Commons platform. Data sharing outside UCSF is restricted and requires a formal data use agreement.

## Data Availability

The UCSF DeID CDW and SFDPH DeID CDW datasets are de-identified clinical relational data repositories available to UCSF researchers through the UCSF Information Commons platform. Data sharing outside UCSF is restricted and requires a formal data use agreement.

## Acknowledgements

The authors thank Claudia Matzko for helpful discussion and Noemie Elhadad, Judy Regensteiner, and Wendy Nilsen for organizing the September 2024 NIH-NSF workshop “Using AI to Better Understand Menopause”, which inspired and motivated this project. The authors acknowledge the support of Google and Apple Machine Learning.

## Author Contributions

I.Y.C. and N.T. conceived of the project idea, and I.Y.C oversaw the project. I.Y.C. and N.T. developed the methodology, and N.T. performed all computations and analyses. R.P. and R.L.G. provided guidance on the cohort creation and data extraction pipeline. J.Z., M.A., and Y.H. verified the analytical methods. All authors discussed the results and contributed to the final manuscript.

## Appendices

**Supplementary Figure S1:**
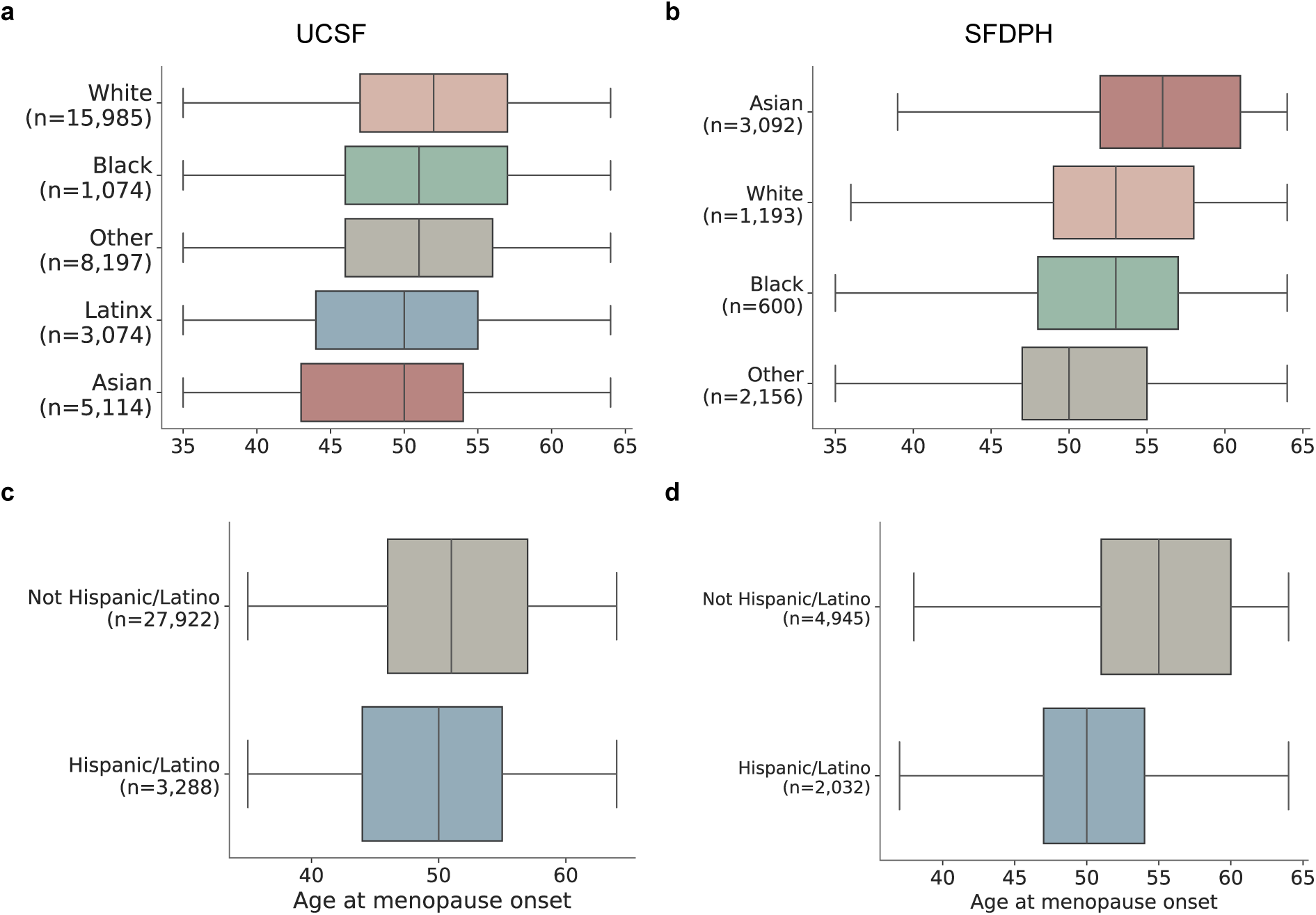
Age at menopause onset by race in the **a.** UCSF and **b.** SFDPH cohorts, and by Hispanic/Latino ethnicity in the **c.** UCSF and **d.** SFDPH cohorts, adjusted for demographic and socioeconomic confounders. Racial differences were site-specific and in opposing directions for Asian women (2.6 years earlier than White at UCSF; 2.8 years later at SFDPH), consistent with ICD-10-based ascertainment bias from differential healthcare access in the safety-net setting. Hispanic/Latino women reached menopause significantly earlier than non-Hispanic/Latino women at both sites (UCSF unadjusted *p* = 1.46 × 10*^−^*^21^; SFDPH unadjusted *p* = 7.77 × 10*^−^*^148^). Latinx race was not recorded separately in the SFDPH dataset, so Hispanic/Latino identity is captured through the ethnicity field.

**Supplementary Figure S2:**
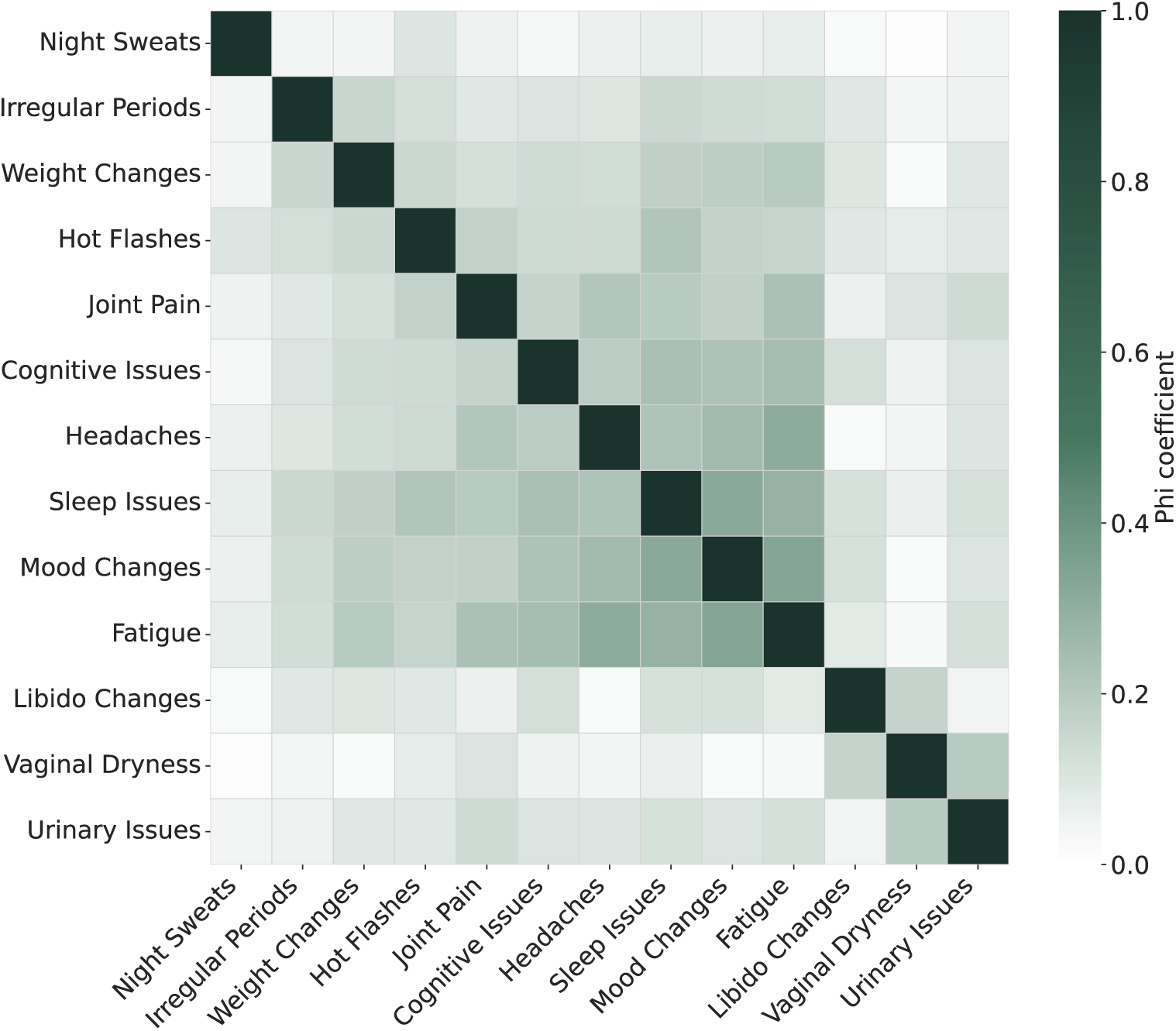
Pairwise co-occurrence of menopause-related symptoms measured by the Phi (*ϕ*) coefficient (darker green indicates stronger positive association). Off-diagonal correlations were uniformly weak (*ϕ* ≤ 0.35). The five strongest associations were concentrated within neuropsychological and somatic symptoms: mood changes–fatigue (*ϕ* = 0.334), sleep issues–mood changes (*ϕ* = 0.318), headaches–fatigue (*ϕ* = 0.308), sleep issues–fatigue (*ϕ* = 0.283), and headaches–mood changes (*ϕ* = 0.250). Vasomotor symptoms (night sweats, hot flashes) and genitourinary symptoms (vaginal dryness, urinary issues) showed little cooccurrence with one another or with other symptom categories.

**Supplementary Figure S3:**
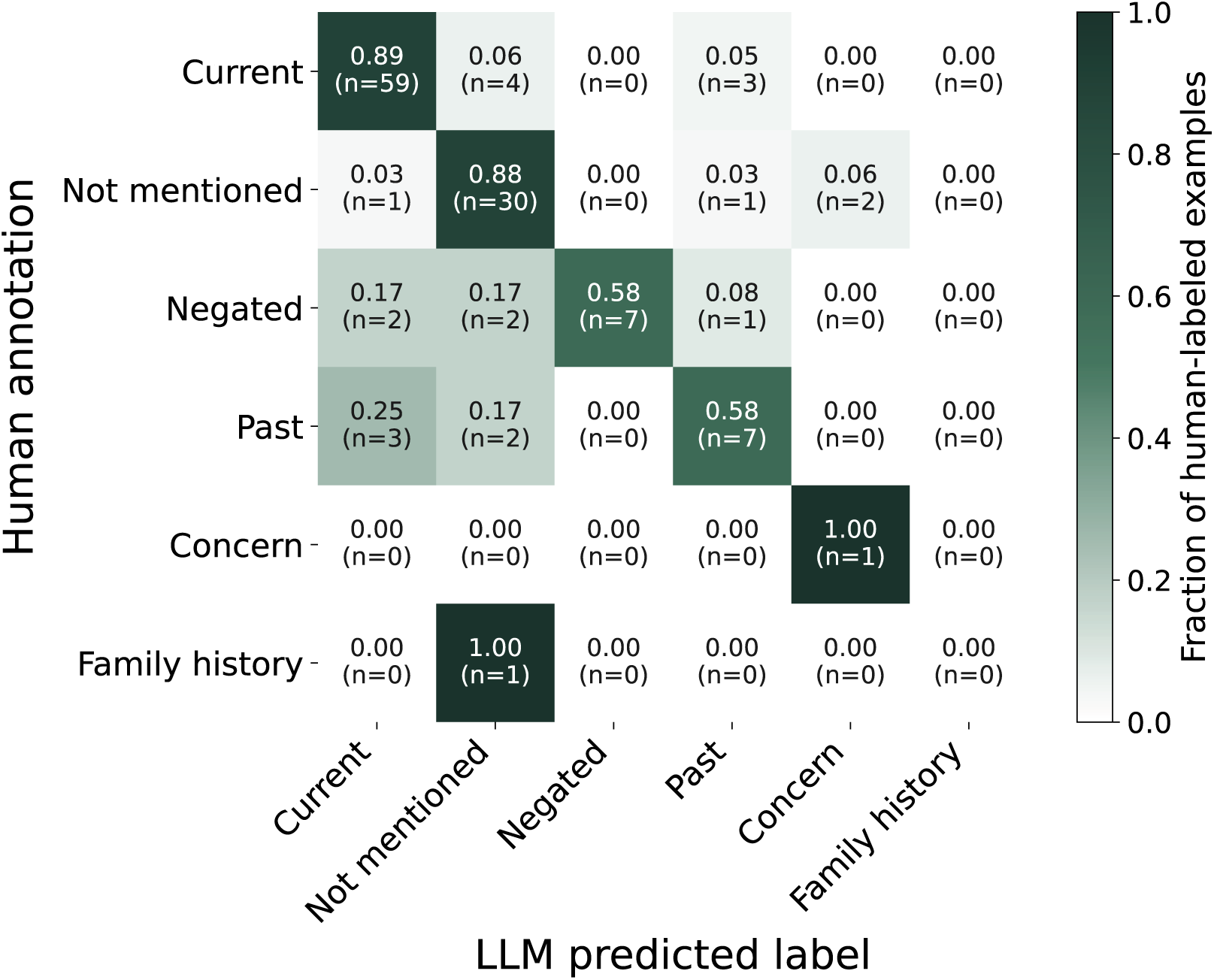
Confusion matrix for LLM symptom context classification validation. Each row represents a human-annotated label, and each column represents the LLM-predicted label. Values are row-normalized so that each cell shows the proportion of human-annotated examples of a given label assigned to each LLM predicted label. Raw counts are shown in parentheses. The LLM achieved the highest accuracy on the ‘current’ label (F1 = 0.89), which drove all symptom prevalence estimates reported in this study, and on ‘not mentioned’ (F1 = 0.80), validating the false positive step of the two-stage pipeline. The lowest performance was observed for rare labels (‘concern’, ‘family history’), which together account for fewer than 3 of 130 annotated examples and do not contribute to prevalence estimates.

**Supplementary Table S1:** Key prior cohort studies of menopausal health outcomes. “Racially diverse” reflects the study’s own reported racial/ethnic composition; studies are flagged as diverse if non-White participants exceeded 20% of the enrolled sample. All cohorts are longitudinal.

| Study (year) |  |  | *N* | Racially diverse | Study design | Age |
| --- | --- | --- | --- | --- | --- | --- |
| UK Biobank [9–11] (2006) |  |  | ~271,791 | No (94.6% White European) | Prospective biobank | 40–69 |
| Nurses’ Health Study [5] (1976) |  |  | 121,700 | No (97% White) | Prospective | 30–55 |
| Nurses’ Health Study II [6] (1989) |  |  | 116,429 | No (92.8% White) | Prospective | 25–42 |
| Women’s Health Initiative [8] (1992) |  |  | 93,676 | No (83.5% White) | Prospective (observational) | 50–79 |
| SWAN [4] (1996) |  |  | 3,302 | Yes | Prospective | 42–52 |
| Multi-Ethnic Study of Atherosclerosis [7] (2000) |  |  | 2,509 | Yes | Prospective | 45–84 |
| UCSF (Ours) |  |  | 33,444 | Yes | Retrospective EHR | 35–64 |
| SFDPH (Ours) |  |  | 7,041 | Yes | Retrospective EHR | 35–64 |

**Supplementary Table S2:** Comparison of demographic and socioeconomic characteristics between the UCSF and SFDPH menopause cohorts. The two cohorts are demographically distinct: UCSF is 47.9% White with a median menopause age of 51 years, while SFDPH is majority Asian (43.9%) with a median menopause age of 54 years and substantially lower rates of MHT use (1.4% vs. 16.0% post-menopause onset), reflecting its safety-net patient population.

| Characteristic | UCSF (*n* = 33,444) | SFDPH (*n* = 7,041) |
| --- | --- | --- |
| *Age at first menopause diagnosis* |  |  |
| Mean (SD) | 50.9 (7.5) | 53.7 (6.2) |
| Median [IQR] | 51 [46–56] | 54 [49–59] |
| Age 35–44 | 7,102 (21.2%) | 451 (6.4%) |
| Age 45–54 | 15,244 (45.6%) | 3,429 (48.7%) |
| Age 55–64 | 11,098 (33.2%) | 3,161 (44.9%) |
| *Race* |  |  |
| White | 16,022 (47.9%) | 1,193 (16.9%) |
| Asian | 5,151 (15.4%) | 3,092 (43.9%) |
| Black | 1,084 (3.2%) | 600 (8.5%) |
| Latinx | 3,031 (9.1%) | 0 (0.0%) |
| Other | 8,156 (24.4%) | 2,156 (30.6%) |
| *Hispanic/Latino ethnicity* |  |  |
| Yes | 3,288 (9.8%) | 2,032 (28.9%) |
| No | 27,922 (83.5%) | 4,945 (70.2%) |
| Unknown/Declined | 2,234 (6.7%) | 64 (0.9%) |
| *Menopausal hormone therapy use* |  |  |
| Any MHT | 9,821 (29.4%) | 213 (3.0%) |
| Post-menopause onset MHT | 5,355 (16.0%) | 98 (1.4%) |
| *Postal code* |  |  |
| 941 | 9,761 (29.2%) | 6,480 (92.0%) |
| 949 | 7,645 (22.9%) | 23 (0.3%) |
| 940 | 3,346 (10.0%) | 219 (3.1%) |
| 945 | 2,946 (8.8%) | 150 (2.1%) |
| Other | 9,746 (29.1%) | 169 (2.4%) |
| *Smoking status* |  |  |
| Never | 22,801 (68.1%) | 3,450 (49.0%) |
| Former | 5,947 (17.8%) | 468 (6.6%) |
| Current | 827 (2.5%) | 429 (6.1%) |
| Unknown/not recorded | 3,869 (11.6%) | 2,694 (38.3%) |
| *BMI* |  |  |
| *N* with BMI | 19,782 (59.1%) | 2,060 (29.3%) |
| Mean (SD) | 25.9 (6.1) | 29.2 (7.1) |
| Median [IQR] | 24.4 [21.7–28.5] | 28.1 [24.3–32.7] |
| *Healthcare utilization* |  |  |
| Mean (SD) | 16.2 (19.1) | 18.7 (15.5) |
| Median [IQR] | 9.3 [4.2–20.7] | 14.6 [9.1–23.1] |
| *Preferred language* |  |  |
| English | 32,094 (96.0%) | 2,477 (35.2%) |
| Spanish | 531 (1.6%) | 1,626 (23.1%) |
| Cantonese/Chinese | 466 (1.4%) | 2,340 (33.2%) |
| Other | 353 (1.0%) | 598 (8.5%) |
| *Marital status* |  |  |
| Married | 20,239 (60.5%) | 3,136 (44.5%) |
| Single | 6,577 (19.6%) | 2,420 (34.4%) |
| Divorced | 2,298 (6.9%) | 943 (13.4%) |
| Widowed | 490 (1.5%) | 300 (4.3%) |
| Other | 3,840 (11.5%) | 242 (3.4%) |

**Supplementary Table S3:** Context classification distribution among keyword-flagged clinical notes for each menopause-related symptom. Percentages represent the proportion of flagged notes assigned to each context label. Family history accounted for *<*0.3% across all symptoms and is omitted.

| Symptom | Notes | Patients | Current | Not Mentioned | Negated | Past | Uncertain | Concern |
| --- | --- | --- | --- | --- | --- | --- | --- | --- |
| Irregular periods | 50,721 | 12,189 | 43.76% | 43.85% | 2.94% | 8.30% | 0.50% | 0.64% |
| Mood changes | 43,151 | 9,826 | 50.51% | 34.25% | 7.14% | 6.10% | 1.17% | 0.82% |
| Fatigue | 19,334 | 6,650 | 70.77% | 12.48% | 7.88% | 6.92% | 0.88% | 1.06% |
| Headaches | 25,629 | 6,607 | 50.01% | 14.80% | 8.40% | 22.69% | 1.99% | 1.95% |
| Sleep issues | 17,729 | 6,102 | 70.80% | 12.51% | 6.67% | 7.89% | 1.44% | 0.68% |
| Vaginal dryness | 10,639 | 4,849 | 61.38% | 29.39% | 3.22% | 2.92% | 1.63% | 1.46% |
| Urinary issues | 13,300 | 4,736 | 67.78% | 14.81% | 5.80% | 8.12% | 1.59% | 1.89% |
| Joint pain | 9,869 | 4,214 | 65.52% | 16.29% | 4.30% | 12.57% | 0.68% | 0.62% |
| Weight changes | 8,305 | 4,124 | 63.17% | 25.29% | 2.66% | 5.05% | 0.54% | 3.30% |
| Hot flashes | 8,188 | 4,030 | 57.73% | 23.82% | 9.99% | 6.17% | 1.16% | 1.14% |
| Cognitive issues | 5,141 | 2,587 | 71.70% | 8.89% | 13.30% | 4.82% | 0.49% | 0.76% |
| Libido changes | 4,104 | 2,160 | 66.28% | 20.54% | 1.02% | 5.80% | 2.53% | 3.83% |
| Night sweats | 270 | 206 | 83.70% | 1.11% | 5.56% | 7.41% | 1.11% | 1.11% |

**Supplementary Table S4:** Hazard ratios (HR) with 95% confidence intervals (CI) and risk-adjusted *p* for UCSF and SFDPH patients when comparing disease risk after menopause onset compared to age-matched men. Risk-adjusted hazard ratio estimates across 12 conditions were highly correlated across cohorts (Pearson *r*=0.80, *p*=1.8×10*^−^*^3^).

| Condition | UCSF ($n = 33,444$) | | SFDPH ($n = 7,041$) | |
| --- | --- | --- | --- | --- |
| | HR (95% CI) | Risk-adjusted $p$ | HR (95% CI) | Risk-adjusted $p$ |
| Osteoporosis | 12.40 (10.55–14.56) | $1.3\times 10^{-206}$ | 10.83 (8.59–13.66) | $3.2\times 10^{-90}$ |
| Atherosclerosis | 3.96 (3.18–4.93) | $1.9\times 10^{-34}$ | 0.65 (0.49–0.85) | 0.0018 |
| Lupus | 2.61 (1.46–4.69) | 0.0013 | 4.37 (1.40–13.61) | 0.011 |
| Rheumatoid arthritis | 2.43 (1.70–3.50) | $1.4\times 10^{-6}$ | 4.09 (2.33–7.17) | $9.0\times 10^{-7}$ |
| Mental/Behavioral disorders | 2.38 (2.22–2.56) | $4.7\times 10^{-126}$ | 2.56 (2.32–2.84) | $4.4\times 10^{-75}$ |
| Alzheimer’s disease | 1.80 (1.04–3.09) | 0.035 | 1.15 (0.39–3.41) | 0.80 |
| Ischemic Heart Disease | 0.87 (0.80–0.93) | $1.5\times 10^{-4}$ | 0.74 (0.62–0.88) | $5.0\times 10^{-4}$ |
| Stroke | 0.79 (0.69–0.91) | 0.0012 | 0.78 (0.64–0.95) | 0.013 |
| Hypertension | 0.69 (0.65–0.74) | $1.4\times 10^{-32}$ | 1.14 (1.03–1.25) | 0.0091 |
| Type 2 diabetes | 0.45 (0.41–0.50) | $3.3\times 10^{-48}$ | 0.73 (0.63–0.84) | $9.0\times 10^{-6}$ |
| Heart failure | 0.33 (0.28–0.39) | $9.6\times 10^{-43}$ | 0.65 (0.53–0.80) | $5.9\times 10^{-5}$ |
| Parkinson’s disease | 0.30 (0.20–0.45) | $4.8\times 10^{-9}$ | 0.38 (0.16–0.90) | 0.028 |

**Supplementary Table S5:** Keyword mapping used to flag menopause-related symptoms in clinical notes. Notes containing any listed keyword (case-insensitive) were flagged for the corresponding symptom category prior to LLM context classification.

| Symptom | Keywords |
| --- | --- |
| Hot flashes | hot flash, hot flush, hotflash, hotflush, heat wave, heatwave, sudden warmth, flushing, vasomotor, warm flash, warm flush, feeling hot, sudden heat, vms, thermal dysregulation, heat intolerance, diaphoresis |
| Night sweats | night sweat, nightsweat, nocturnal sweat, sleep sweat, night time sweat, nighttime sweat, sweating at night, wake up sweating, sweat while sleeping, drenching sweat, nocturnal diaphoresis, sweats at night |
| Irregular periods | irregular period, irregular cycle, irregular menses, irregular menstrual, missed period, skipped period, skipping period, cycle change, menstrual change, period change, cycle abnormal, abnormal period, erratic period, unpredictable period, amenorrhea, oligomenorrhea, menstrual irregularity, menopausal transition |
| Vaginal dryness | vaginal dry, vaginal atrophy, vulvar dry, vulvovaginal dry, vaginal lubrication, lack of lubrication, atrophic vaginitis, dyspareunia, painful intercourse, painful sex, discomfort during sex, vaginal discomfort, vaginal irritation, vaginal burn, gsa, gsm, vva, genitourinary syndrome, vulvovaginal atrophy, sexual pain |
| Mood changes | mood swing, mood swings, irritability, irritable, anxiety, anxious, depression, depressed, mood change, emotional lability, mood disorder, weepy, agitated, overwhelmed, emotional instability, labile mood, mood lability, perimenopausal mood |
| Sleep issues | insomnia, sleep disturbance, difficulty sleeping, trouble sleeping, sleep problem, restless sleep, poor sleep, sleep disorder, can't sleep, cannot sleep, difficulty falling asleep, frequent waking, sleep disruption, unrefreshing sleep, sleep fragmentation, early morning awakening, middle insomnia |
| Cognitive issues | brain fog, memory loss, memory problem, forgetful, forgetfulness, concentration difficult, difficulty concentrating, can't focus, cognitive decline, mental fog, fuzzy thinking, focus issue, attention problem, word finding, cognitive complaint, subjective cognitive, menopausal brain fog |
| Joint pain | joint pain, joint ache, arthralgia, muscle ache, stiffness, joint stiffness, body ache, aching joints, muscle pain, myalgia, joint soreness |
| Weight changes | weight gain, weight increase, gained weight, gaining weight, abdominal weight, belly fat, midsection weight, metabolism slow, slow metabolism, metabolic change, difficulty losing weight, hard to lose weight, can't lose weight, visceral adiposity, central adiposity |
| Fatigue | fatigue, fatigued, tired, tiredness, exhausted, exhaustion, low energy, lack of energy, no energy, lethargy, lethargic, worn out, sluggish |
| Headaches | headache, headaches, migraine, migraines, head pain, cephalgia, tension headache |
| Urinary issues | urinary frequency, frequent urination, urinary urgency, incontinence, incontinent, bladder issue, bladder problem, leaking urine, bladder control, stress incontinence, urge incontinence |
| Libido changes | decreased libido, low libido, libido, sex drive, sexual dysfunction, loss of interest, sexual interest, sexual desire, hypoactive sexual desire, sexual avoidance |

